# Hearing screening beyond the clinic: Childhood cancer survivors’ perspectives on a novel hearing screening program

**DOI:** 10.1101/2025.02.27.25322837

**Authors:** Philippa Jörger, Carina Nigg, Leonie Schreck, Christina Schindera, Katharina Fessler, Nicolas Waespe, Zuzana Tomášiková, Marc Ansari, Annika Frahsa, Claudia Kuehni

**Author notes:** Corresponding author: Correspondence to: Claudia E. Kuehni, Prof. MD; Institute of Social and Preventive Medicine, University of Bern, Mittelstrasse 43, 3012 Bern, Switzerland. These authors contributed equally (Annika Frahsa, Claudia Kuehni).

## Abstract

**Purpose:** Childhood cancer survivors (CCS) are at risk for late effects including hearing loss due to ototoxic treatments. It is crucial to detect late effects like hearing loss early, but many adult CCS do not attend recommended follow-up care due to barriers such as accessibility and emotional concerns associated with revisiting medical facilities. To address those barriers, the HEAR-study piloted a new hearing screening program leveraging the extensive network of hearing aid shops across Switzerland. This study explored perspectives of CCS on this novel screening program.

**Methods:** We conducted semistructured interviews with 29 participants who completed pure-tone audiometric screening at a hearing aid shop as part of the HEAR-study. We used thematic analysis of interview transcripts, supported by MAXQDA for data analysis.

**Results:** Thematic analysis revealed two key themes: First, participants appreciated the program’s practicality, highlighting its efficiency and ease of integration into daily life. However, some noted concerns about the hearing test being an additional appointment. Some preferred centrally organized follow-up care, where different examinations are done at the same location within the same day. Second, participants valued the personal and approachable environment in hearing aid shops as a relaxed alternative to medical facilities, but some participants preferred hearing tests as part of an appointment with physicians, valuing immediate advice and contextual knowledge, especially in case hearing loss was found.

**Conclusions:** From the perspectives of CCS, this screening program shows promise as a practicable, accessible way to evaluate hearing after childhood cancer. Further evaluation from the perspectives of health care providers is needed to comprehensibly assess its feasibility.

**Trial registration:** ClinicalTrials.gov “NCT06036407”, Registration date: 28.8.2023

## Introduction

Childhood cancer survivors (CCS) are at risk for cancer- and treatment-related long-term health conditions that can occur years after treatment completion [1]. Potential late effects include hearing loss caused by ototoxic treatments such as cranial radiation and platinum chemotherapy, whereby 10% of all CCS develop hearing loss [2]. Unrecognized, even mild hearing loss can negatively affect communication, education, employment and quality of life, which can be prevented with timely interventions like hearing aids or communication strategies [3–6]. To detect such late effects early and take appropriate measures, risk-based follow-up care is crucial [7–10]. Follow-up care refers to scheduled consultations aimed at monitoring for cancer recurrence and potential late effects of treatment.

Despite the importance of follow-up care, a significant proportion of adult CCS do not attend care as recommended [11–16]. In Switzerland only half of participating CCS are reported to have attended any follow-up care [15], while a study from France found even lower follow-up attendance of 33% [11]. In North America and Europe, follow-up care for pediatric and adolescent CCS is initially organized within the pediatric oncology outpatient clinic where survivors received cancer treatment [17–19]. Survivors and their parents discuss results of follow-up examinations with pediatric oncologists and specialists in potentially involved organ systems. Once survivors reach adulthood, CCS transition to adult follow-up care, which is either carried out by a primary care physician, an adult oncologist, or set up within a structured long-term follow-up care (LTFU) program. LTFU programs consist of a multidisciplinary team lead by an internal medicine physician and include specialized nurses, different medical disciplines, social workers, and psychologists [17–19]. In Switzerland, structured LTFU programs for risk-adapted follow-up care have been established in several centers, but they are resource intensive and currently only available for a small proportion of CCS. Previous studies suggest various barriers to follow-up attendance that include lack of knowledge about potential late effects, lack of knowledge of follow-up care possibilities [16,20], long distances to follow-up care appointments [14,16], costs associated with medical appointments [21,22], and emotional barriers related to the cancer experience and fear of pathological findings [23,24].

To address some of these barriers, the HEAR-study piloted a new screening program designed to provide a low-threshold option to screen for hearing loss after childhood cancer [25]: In contrast to customary follow-up appointments, the program leverages the extensive network of hearing aid shops distributed throughout Switzerland to offer an easily accessible, alternative site at which survivors may undergo high quality hearing tests that require minimal time, effort, and financial resources from individuals. In Switzerland, hearing aid shops are retail outlets that sell hearing aids and offer hearing tests free of charge to all individuals who wish to check their hearing. By taking place outside a hospital or doctor’s office, this screening program may encourage follow-up attendance by CCS who might otherwise be reluctant to revisit medical facilities.

The overall aim of the HEAR-study is to evaluate the feasibility, limitations, and potential for improvement of this screening program. We therefore interviewed participating CCS who completed an audiometric pure-tone hearing test in a hearing aid shop to learn and interpret their perspectives on such screening, which are crucial to further developing the program.

## Methods

### Study design

This study is part of the HEAR-study, a health service research project set up in 2021 in Switzerland. Details of the HEAR-study are published elsewhere [25]. The HEAR-study piloted a new screening program for hearing loss after childhood cancer. Briefly, as part of the hearing screening program, we invited eligible CCS (N=1,604) to complete pure-tone audiometric screening at hearing aid shops across Switzerland belonging to one company. Participants arranged an appointment independently at a shop of their choice. Audiometry included air conduction testing from 125 to 8,000 Hz; bone conduction (250-4,000 Hz) was added if hearing loss exceeded 25 dB. The hearing tests were conducted by hearing aid acousticians employed at the hearing aid shops. Acousticians in Switzerland complete a three-year vocational training, earning a federally recognized certification as hearing aid technician. Acousticians provided participants with the results both verbally and as a printout, encouraging them to consult their physician if results were borderline or pathological. For ethical reasons, acousticians were not informed about participants’ medical histories. While the shops have high quality equipment for audiometric screening, they do not have fully soundproof chambers. This manuscript presents qualitative data from interviews with HEAR-study participants that provide insights into their experiences, perceived barriers, and reasons for favoring or not favoring this screening program. The Ethics Committee of the Canton of Bern approved the Childhood Cancer Registry (ChCR) and the HEAR-study (166/2014; 2021-01624). We used the Consolidated criteria for Reporting Qualitative research checklist (COREQ) as guidance for reporting [26].

### Participant selection

CCS eligible for the HEAR-study were identified through the Swiss ChCR. The ChCR is a national registry of Swiss residents diagnosed with leukemias, lymphomas, central nervous system (CNS) tumors, malignant solid tumors, or Langerhans cell histiocytosis before 20 years of age and since 1976 [27,28]. We obtained cancer- and treatment-relevant information from the ChCR.

All registered CCS who were ≥18 years at study start and who were at increased risk for hearing loss due to their treatment—any chemotherapy and/or radiation to head, neck or spine—were eligible for the HEAR-study. We distinguished between two risk groups, high risk, treated with known ototoxic treatments including cranial radiation ≥30 Gray or cisplatin/carboplatin, and standard risk, treated with any other chemotherapy or radiation to head, neck or spine with any dose. We excluded CCS with a correspondence language other than German or French due to limited resources, those for whom we had no current postal address, and those who had recently been contacted for another study (January-July 2022) so as not to overburden them.

Of 1604 invited, 319 completed a hearing test after providing baseline sociodemographic data. Participants were then sent a follow-up survey to indicate interest in an interview; 296 returned this survey, and 169 expressed interest. We used purposive sampling to select a diverse sample of interview participants regarding current age, sex, education, corresponding language (German/French), and risk group for hearing loss.

### Data collection

Invited interview participants could choose face-to-face interviews to be conducted online or in person at a location of their choice. We informed participants about the goals of the study, explained data anonymity, and collected written consent before the interview. We used a semistructured interview guide that focused on participants’ experiences at the hearing aid shops and advantages and disadvantages in conducting hearing tests as part of follow-up care at hearing aid shops compared to medical institutions (e.g., hospitals). The interview guide used was developed for this study (supplementary table S1). We piloted the interview guide with study team members and iteratively refined it based on feedback and insights from initial interviews with study participants; changes improved question clarity, sequence, and wording.

We planned to interview between 20-30 participants for practical reasons and based on the concept of information power, considering our narrow study objective and specific sample [29]. After conducting most of the interviews and simultaneous data analysis, we decided to invite 30 participants to capture a sufficient large number of participants. One person declined, and after completing and coding 29 interviews, we determined that we collected sufficient data to meaningfully explore the diverse perspectives of CCS. We interviewed participants from October 2022 to April 2023, with the main author (PJ, female, PhD student) conducting 24 interviews in German and a trained research assistant (female) conducting 5 interviews in French, with PJ providing support in the event of ambiguities or questions arising from participants. Quotes in this manuscript were translated from German or French to English and edited for language and clarity (supplementary table S2).

### Data analysis

All interviews were recorded (mean length: 31min; range 17-53 min) and, if conducted in French, translated into German by a bilingual research assistant familiar with the study context. During the interviews, the interviewer made notes on interesting aspects discussed and the overall impression of the interviewee for later consideration during data analysis. Two research assistants (female) did verbatim transcriptions into Microsoft Word before importing into MAXQDA 2022 (VERBI Software 2021. Consult. Sozialforschung GmbH, Berlin, Germany) for detailed analysis and verified all transcripts against the audio recordings. Braun and Clarke’s (2021) approach to reflexive thematic analysis served as guiding framework for data analysis [30], with the research team positioned in a critical realist perspective.

Two researchers (PJ and KF), both trained in qualitative research methods, inductively coded a subsample (20%) of interviews independently, guided by the research question. Both discussed their coding from the subsample before agreeing upon a similar coding scheme, which they then discussed with the rest of the research team. This included an established expert in qualitative research (AF, female, professor in community health).

After refining the coding scheme with the study team, both researchers independently coded the remaining transcripts. One researcher (PJ) then aligned the codes with existing themes and subthemes and identified new ones. We produced a brief description of each theme to clarify the content, check for overlaps between themes, and establish final themes and subthemes to present findings clearly and coherently within the narrative. During the coding process we discussed themes/subthemes regularly within the whole research team (eight women, two men, with expertise in pediatrics, pediatric oncology, childhood cancer survivorship, social and preventive medicine, community health) to check for trustworthiness of the findings. The coders kept reflexive diaries, documenting changes in codes and themes, along with discussions and decisions made throughout the process, and personal thoughts and potential preconceptions. We analyzed the data on a semantic level and identified themes that captured the diverse range of experiences and perspectives of participants with the screening program.

## Results

### Sample characteristics

Twenty-nine people (15 women) with a median age of 36 years at study start and median age of 12 years at diagnosis participated in the study. They had attended appointments at 15 different hearing aid shops across Switzerland. Eleven participants were in the high-risk group for hearing loss (cf. further information Table 1). Fifteen interviews took place online, eight were conducted in a meeting room at the University of Bern, while four were conducted at participants’ homes and one each in another university meeting room and a workplace.

**Table 1:** Participant demographic information and disease characteristics. (N=29)

| **Characteristics** | **n (%)** |
| --- | --- |
| Sex |  |
| Male | 14 (48%) |
| Female | 15 (52%) |
| Age at study in years<br>(median, range) | 36 (18-56) |
| Age at diagnosis in years<br>(median, range) | 12 (0-17) |
| Diagnosis year |  |
| <1986 | 3 (10%) |
| 1986-1995 | 8 (28%) |
| 1996-2005 | 12 (41%) |
| >2005 | 6 (21%) |
| Cancer diagnosis |  |
| Leukemias | 5 (17%) |
| Lymphoma | 8 (28%) |
| CNS | 2 (7%) |
| Neuroblastoma | 3 (10%) |
| Malignant bone tumor | 5 (17%) |
| Other<sup>a</sup> | 6 (21%) |
| Risk group |  |
| High risk<sup>b</sup> | 11 (38%) |
| Standard risk | 18 (62%) |
| Correspondence language |  |
| German | 24 (83%) |
| French | 5 (17%) |
| Education level |  |
| Primary or secondary | 16 (55%) |
| Tertiary | 13 (45%) |
| Living area<sup>c</sup> |  |
| Urban | 18 (62%) |
| Intermediate | 7 (24%) |
| *Rural* | 4 (14%) |
| *Still visiting regular follow-up care* |  |
| Yes | 16 (55%) |
| No | 13 (45%) |
| *Self-reported hearing loss after the hearing test* |  |
| Yes | 13 (45%) |
| No | 16 (55%) |

<sup>a</sup>Others include: Soft tissue sarcoma, Hepatic tumor,
Retinoblastoma, Renal tumor, Germ cell tumor.
<sup>b</sup>Ototoxic treatments according to International Guideline
Harmonization Group guidelines [31]; cisplatin, carboplatin, or cranial
radiation $\geq 30$ Gray.
<sup>c</sup>According to the Swiss Federal Office for Statistics; Typology of
municipalities and urban-rural typology [32].

### Themes

Two main themes represent key perspectives of CCS on the feasibility of the program (Table 2). The first theme focused on practicality, which participants identified as a crucial aspect of follow-up care appointments. The second theme concerned the different circumstances of hearing tests in a hearing aid shop compared to a medical institution (e.g., hospital) and their advantages and disadvantages. In general, participants reported positive experiences with the hearing test at the hearing aid shops. Only a few reported negative experiences such as feeling tense during the test or experiencing slight discomfort during the examination.

**Table 2:**
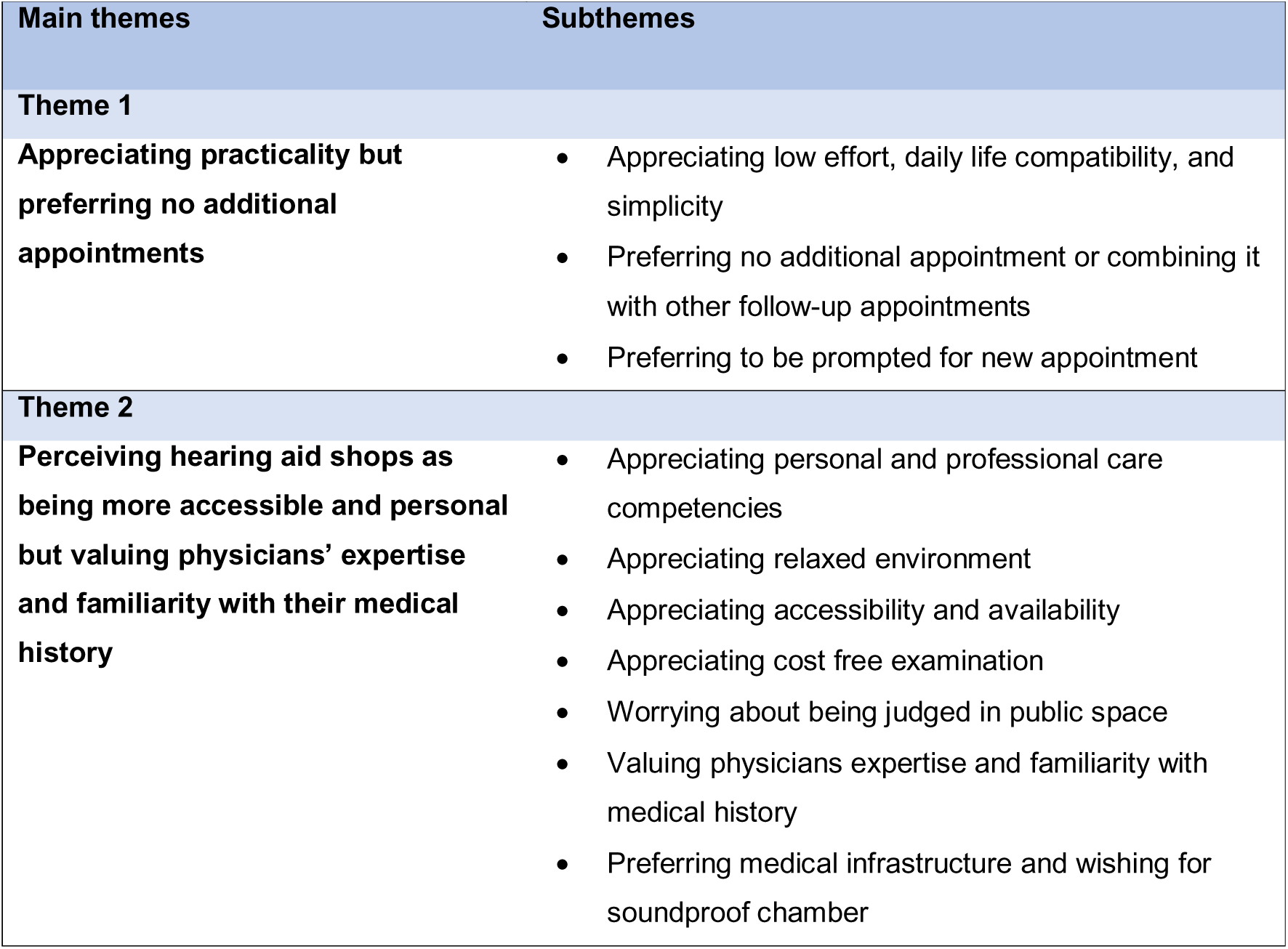
Overview of main themes and corresponding subthemes.

| Main themes | Subthemes |
| --- | --- |
| **Theme 1** |  |
| **Appreciating practicality but preferring no additional appointments** | • Appreciating low effort, daily life compatibility, and simplicity<br>• Preferring no additional appointment or combining it with other follow-up appointments<br>• Preferring to be prompted for new appointment |
| **Theme 2** |  |
| **Perceiving hearing aid shops as being more accessible and personal but valuing physicians' expertise and familiarity with their medical history** | • Appreciating personal and professional care competencies<br>• Appreciating relaxed environment<br>• Appreciating accessibility and availability<br>• Appreciating cost free examination<br>• Worrying about being judged in public space<br>• Valuing physicians expertise and familiarity with medical history<br>• Preferring medical infrastructure and wishing for soundproof chamber |

### Appreciating practicality but preferring no additional appointments

Participants prioritized efficiency and practicality in managing follow-up care appointments such as a hearing test. Participants’ experiences at the hearing aid shops aligned with their wish for practicality. They described procedures at the hearing aid shops as efficient and uncomplicated: scheduling the appointment was easy, the examination was carried out swiftly, and it was overall, as a few participants said, “no big deal.” They also liked that no prior consultation or long waiting times were needed, compared to attending doctors’ appointments. Participants perceived the test as compatible with activities of daily life: they appreciated the options to have the hearing test before starting work, during their lunch break, or while taking care of their children. Some compared the effort of the hearing test to other appointments in daily life such as getting a haircut or buying shoes, and reported they completed their daily activities around the visit.

> *“I was able to make the appointment in the morning so that it wasn’t in the middle of the day. Then I could do it before work. […] They were… well, it was convenient.” 30-39-year-old woman*

A few participants experienced the visit to the hearing aid shop as less practical and compatible with everyday life. For them, the effort was higher when the hearing aid shop they visited was not nearby. In these cases, participants visited the hearing aid shop on days off work such as weekends.

Several survivors stated they preferred not having any additional appointments in their schedule or preferred a structured LTFU program to avoid additional effort, i.e. having to visit different locations on different days. Some also mentioned that in the event of an abnormal result they would have to attend a doctor’s appointment anyway, which would mean even more effort for them.

> *“If you’re having a follow-up check-up with the oncologist anyway […] and you can do it all at the same time: a lab appointment, an X-ray appointment and a hearing test at the same place on the same date, most people would certainly appreciate that. […] Then you don’t have to drive to this place, that place and yet another place. You could do it all in one go.”*

### 30-39-year-old woman

Some individuals expressed that the responsibility and organizational effort of scheduling follow-up appointments was burdensome, highlighting the significance of reminders and being invited to appointments. One person proposed a centralized system that would provide an overview of all their follow-up appointments, that would remind them of appointments or even help with scheduling.

### Perceiving hearing aid shops as being more accessible and personal but valuing physicians’ expertise and familiarity with their health history

Participants were content with interpersonal care (i.e. how they were treated) and professional competencies of acousticians at the hearing aid shops. The majority particularly appreciated the thorough explanations and information they received during the visit about hearing test procedures, test results, and hearing-related topics raised during the test. Only a few participants were left with open questions about specific topics (e.g., hearing hypersensitivity, tinnitus).

> *“I was warmly welcomed and looked after. They also told me a bit about what they were doing and why, and explained how the hearing [physiology] works. They also explained possible hearing impairments […] and how the test is conducted. I really felt very comfortable there and if I had had any questions, I would certainly have been able to ask them, but they were actually answered by the person during the process.” 30-39-year-old man*

Several participants perceived the atmosphere of the hearing aid shops as more relaxed and personal than in a medical facility, which they valued. Some participants appreciated that the acoustician was on their level and that there was more interaction compared to a medical doctor. One person mentioned that this made them feel more comfortable asking questions, indicating that there was a lower barrier to engage with acousticians compared to medical doctors, who were perceived as being less approachable.

> *“It was definitely quicker and, for me personally, a better experience than at the ENT specialist. […] It was more conversational, more interactive for me. I learned more from this test than I did from the one at the doctor.”*

### 18-29-year-old man

They considered this relaxed environment as advantageous compared to medical facilities, considering some might have a negative association with medical facilities related to their cancer history.

> *“A hearing aid shop is low threshold. It’s not: ‘Ah, I’m going back to the hospital now’. Depending on the situation, you might not have the best associations with hospitals. And if you’re just like anyone else who goes there for counseling, it’s actually pleasant. It’s not the feeling of being a patient, but simply a customer.” 30-39-year-old woman*

Survivors appreciated that the hearing aid shops were accessible, not just in terms of the geographic location, but also in being approachable and available without referral or cost.

> *“It’s a place where I can go again at any time. Let’s say I want to know something more, or I still want to have this photo [otoscopy]. I can just write an email or call. The shop is always available. I mean these specialists [doctors], just getting an appointment at all… they are somehow so far away. Not so close. Not so easy to reach. And that seems to me to be a clear advantage [of the hearing aid shops].” 40-49-year-old woman*

A few participants appreciated the test being free of cost. They emphasized that cost of examinations was an important barrier to follow-up care appointments. While health insurance is mandatory in Switzerland, there is a minimum deductible that patients must pay themselves for examinations.

One participant noted a downside of the hearing aid shop’s open and approachable layout: being in a public space might be uncomfortable for survivors with visible side or late effects of their therapy. The community-based environment might lead to them feeling judged by people who are not physicians.

> *“I see a disadvantage [of the hearing aid shops] for someone who […] still has visible sequelae of the illness [later he mentioned hair loss as an example]. That may really hold someone back from going to a public place.”*

### 40-49-year-old man

Although many participants felt well looked after of at the hearing aid shops, some still perceived physicians as more knowledgeable and trustworthy. They argued that physicians had extensive education and contextual knowledge that is important for conducting examinations and interpreting results in relation to their cancer history.

> *“[Examinations in hearing aid stores] have no medical components. They don’t have a doctor to interpret [the results]. Not to question the competence of a hearing aid acoustician, but they don’t have medical staff who know my diagnosis and can reliably assess [the hearing]. And that’s why I would say the hearing aid shop is certainly quicker and better for people who just want a rough assessment of how well they can hear. But it’s not suitable for people who really need medical follow-up treatment and follow-up checks like I do, because they don’t meet all the requirements. Even if it might be a bit more practical.”*

### 18-29-year-old man

Some participants reflected on the importance of trust, familiarity, and continuity in follow-up care. A few individuals specifically mentioned that they preferred “receiving bad results” from a physician. Some emphasized the importance of always being examined by the same people for several reasons. They found it burdensome to repeatedly educate new health care professionals about their cancer history, worried about information loss if different health care professionals were involved in the same examination, and saw value in having an established relationship with health care staff, especially for sensitive issues. In relation to this, some participants wished for a person with an overview of their medical history and whom they could contact in case of questions.

> *“I think the advantage, for example, with a doctor or at the hospital is that they might… know my medical history and maybe be able to say quickly: “Yes, that could be it. That’s possible.” […] Or they might be able to specifically relate it back to my cancer diagnosis. […] That they might also be able to respond and check if something’s wrong. […] Whereas [name of hearing aid company] […] doesn’t know about my cancer history and just looks at it more superficially.”*

### 18-29-year-old woman

Nevertheless, many agreed that hearing aid shops could serve as a useful opportunity for initial hearing screening, with referral to a physician in case of unusual findings. For several, integrating the hearing test in everyday life was still more important than the type of institution.

> *“If I had to do such tests more often, the most decisive factor would probably be where it is easiest to access… Locally. So it would be easy to integrate into everyday life. I wouldn’t care whether it was a clinic or a hearing aid shop. Where I live, I would probably rather go to the [hospital name]. On the other hand, you wait longer in the clinic. At [hearing aid company name], they "timed" it nicely that it was just my turn. That is a further aspect. So above all: what is easiest to integrate into everyday life?”*

### 30-39-year-old woman

Some participants expressed concerns about the commercial nature of hearing aid shops, believing that the acousticians performing the tests were motivated by the potential for a sale. A few were worried that acousticians might try to sell them products that they didn’t need or want. Final comments concern the professionalism of the hearing test infrastructure. Some perceived the infrastructure not to be as professional as in a medical setting. Mainly, they missed a soundproof chamber for the test, and some were distracted by background noise.

Table 3 summarizes some of the advantages and disadvantages of screening hearing in hearing aid shops that were mentioned by participants.

- Accessible and easily available
- Time efficient and convenient
- Personal and approachable environment
- No individual costs for CCS
- Suitable for initial screening when likelihood of hearing loss is low
- More comfortable, non-medical environment
- Burden of additional appointment
- Lack of contextual knowledge (lack knowledge on medical history)
- Lack of immediate consultation with a physician in case of abnormal findings
- Acousticians perceived as less knowledgeable
- No soundproof chamber

**Table 3:**
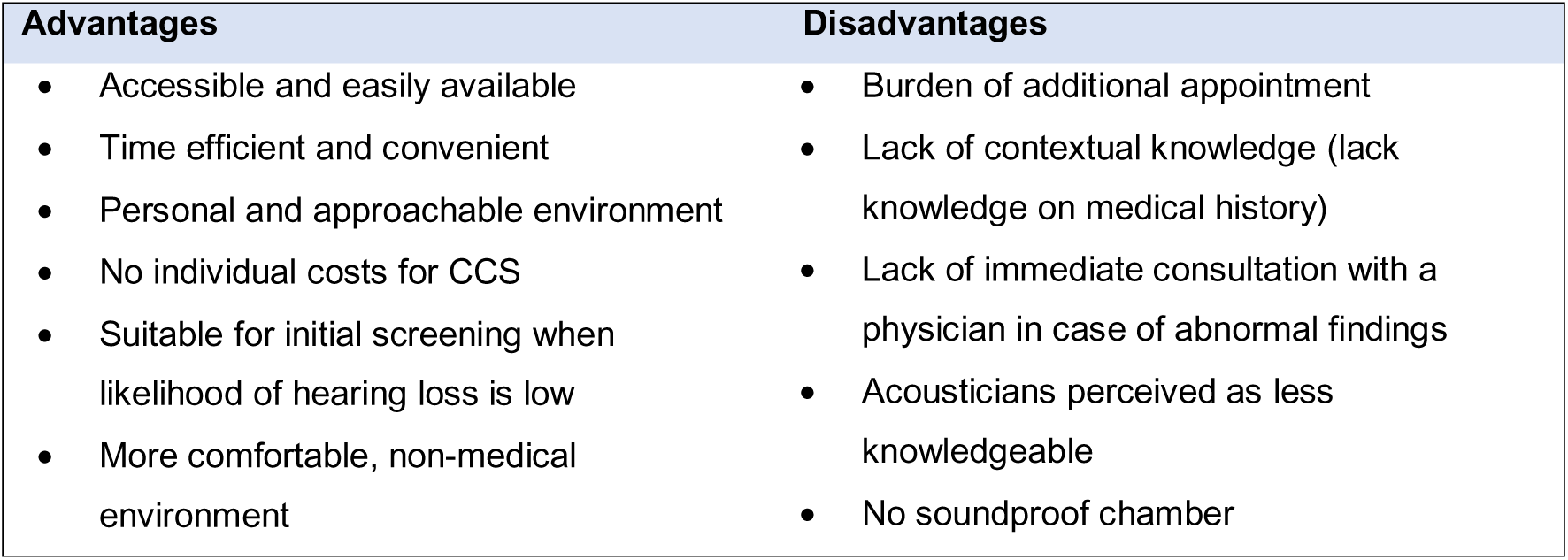
Possible advantages and disadvantages of screening for hearing loss in hearing aid shops mentioned by childhood cancer survivors.

## Discussion

Participants reported many positive experiences with the screening program based in hearing aid shops as proposed within the HEAR-study. They appreciated the practicality and accessibility of hearing aid shops, regarding the shops as a convenient venue for screening. Visits were perceived as relaxed and personal. Participants’ overall concerns included the burden of organizing and attending multiple appointments at different institutions, as well as the perception that physicians are better suited to interpret hearing test results in the context of their medical history.

Practicality and local availability were important for participants when attending follow-up appointments like hearing tests. These HEAR-study findings confirm those of previous studies on barriers to follow-up care showing that local accessibility facilitates participation, while living in remote areas can pose a barrier to accessing care [11,16]. Aligning with reports of adult CCS in previous qualitative studies on follow-up care [20,33], we found that participants value structured LTFU programs: they are reluctant to attend multiple appointments in different institutions due to organizational burden and the effort associated with traveling to different locations. However, when structured LTFU programs are not feasible for all CCS, whether due to limited admission capacity or geographical barriers, the here proposed screening program could provide a practical and straightforward option to screen for selected late effects, such as hearing loss. Participants in the HEAR- study highlighted the practicality and simplicity of their visits at the hearing aid shops.

Reducing the need for travel and simplifying the appointment process could improve adherence to recommended follow-up care. To implement any such program, a robust organizational framework should be established, and CCS should be adequately supported in managing their follow-up appointments. The Survivorship Passport or Passport for Care, electronic treatment summaries including individualized recommendations for follow-up, could guide organization of different appointments within follow-up care [34,35]. Discussions with other stakeholders including pediatric oncologists will shed further light on how to best implement such a program.

Previous studies on preferences for and barriers to follow-up care report that adult CCS value personal care and competent staff during examinations [14,24], which we also found in our study. Most participants felt well cared for and professionally consulted during their visits. Participants appreciated the approachability of staff who made it easy for CCS to engage with acousticians during and after the hearing test. The relaxed atmosphere made the experience more comfortable for some CCS, which is particularly important given psychological burden often associated with follow-up visits. This burden, discussed in our own and other studies on preferences and barriers to follow-up care among adult CCS [16,23,24,33], can prevent CCS from returning to hospitals or healthcare providers; a welcoming environment is crucial for encouraging attendance to follow-up care. CCS also valued the hearing test being free of charge, which aligns with previous findings that highlight costs as a considerable barrier to accessing healthcare [36]. Some participants perceived acousticians as less knowledgeable and preferred physicians who could interpret the results of the hearing test in the context of their cancer history. This aligns with findings from earlier research that highlight survivors’ apprehensions about new healthcare providers lacking an understanding of their medical history and unique healthcare needs [24,37,38].

That said, regardless of whether the hearing test is performed in a hearing aid store or at a medical facility, abnormal or inconclusive results in any case require follow-up consultation with a physician. In a setting in which the likelihood of a pathological finding is relatively low (as in our study, with an expected prevalence of 10%) [2], consultation with a physician need not take place in the majority of cases. Implemented more broadly, such a program should be set up so that only people with a pathological finding would then schedule a medical appointment with a physician. This approach could reduce physician workload and thus help alleviate the increasing shortage of doctors in Switzerland [39]. Many participants agreed that this screening approach, when results are likely to be within a normal range, could be used for initial screening.

## Strengths and limitations

This study demonstrates the potential feasibility of a hearing screening program conducted at hearing aid shops from the perspectives of CCS. Interviewing CCS allowed us to gain insights into their experiences with this program, which are essential to further develop and implement it. By using a semantic approach, we attempted to represent participants’ voices as accurately as possible. It is important to recognize that CCS are a diverse group with varying needs and preferences regarding follow-up care. While some may benefit from such a screening option, others may prefer follow-up care solely in clinical settings. The program could be a complementary option available to CCS so that they can choose the setting that best aligns with their needs. Although we used a thorough coding process with multiple rounds of discussions to ensure reliability, there are always different ways to interpret and code data due to the subjective nature of qualitative analysis. Our study recruited participants from the overall HEAR study and includes only their perceptions. We cannot report on the thoughts or perceptions of survivors who were not tested at a hearing aid shop.

## Conclusion

This study examined the potential of a new hearing screening program designed to provide CCS with a low-threshold alternative to screenings in formal medical settings. Its findings suggest that such a program based in hearing aid shops seems feasible and could facilitate greater participation in follow-up care, particularly among CCS outside structured LTFU who prioritize accessibility, personal atmosphere or fear medical institutions. Informed by survivors’ perspectives, the next step is to explore program feasibility from the perspectives of other stakeholders: health care providers including pediatric oncologists and ear, nose, and throat specialists, and the hearing aid company. This will allow us to work further on exploring how such a program could support decentralized follow-up care in Switzerland.

## Data Availability

The datasets generated or analyzed during this study are stored on secured servers of the Institute of Social and Preventive Medicine at the University of Bern. Individual-level, fully anonymized, sensitive data can only be made available for researchers who fulfil the respective legal requirements.

## List of Abbreviations

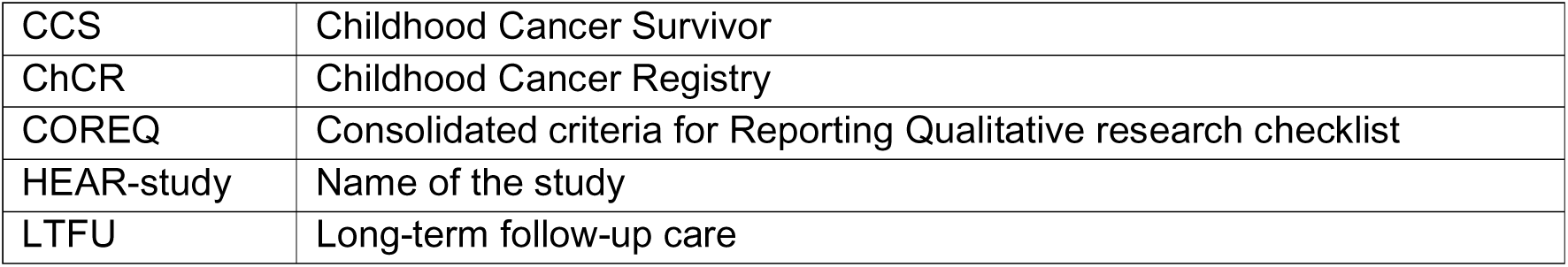

|  |  |
| --- | --- |
| CCS | Childhood Cancer Survivor |
| ChCR | Childhood Cancer Registry |
| COREQ | Consolidated criteria for Reporting Qualitative research checklist |
| HEAR-study | Name of the study |
| LTFU | Long-term follow-up care |

## Declarations

### Ethics approval

All procedures performed in studies involving human participants were in accordance with the ethical standards of the institutional and/or national research committee (Ethics Committee of the Canton of Bern, Switzerland, KEK-BE 2021-01624) and with the 1964 Helsinki declaration and its later amendments or comparable ethical standards. Informed consent to participate was obtained from all individual participants included in the study.

### Consent for publication

Not applicable.

### Availability of data and materials

Research data are not shared due to ethical reasons. Requests of data from the Childhood Cancer Registry must be directed to the Childhood Cancer Registry of Switzerland (https://www.childhoodcancerregistry.ch).

### Author contributions

CK, MA and NW contributed to the study conception and design. Data collection and analysis were performed by PJ and KF and critically assessed by CK and AF. All authors participated in discussions regarding the analysis and interpretation of the data. The first draft of the manuscript was written by PJ, CN, and LS, and all authors commented on previous versions of the manuscript. All authors read and approved the final manuscript.

## Acknowledgements

We thank the study team of the Childhood Cancer Research Group, Institute of Social and Preventive Medicine, University of Bern, the Swiss Childhood Cancer Survivor Study, and the team of the Swiss Childhood Cancer Registry. We thank Leah Weber for her valuable work. We thank all childhood cancer survivors participating in the study and for the interesting discussions. We also thank Christopher Ritter for editorial assistance.

