## Supplementary Material for "Hearing screening beyond the clinic: Childhood cancer survivors’ perspectives on a novel hearing screening program"

### Supplementary TABLE S1 Semistructured interview guide (translated from German to English).

Note: The interview used in our study was developed for this study and has not previously been published elsewhere. Main questions were further explored by follow-up questions.

| Introduction | Interviewer introduced herself briefly and explained again the aims of the HEAR-study and the procedures of the interview. |
| --- | --- |
| Introductory questions | Why don't you start by telling me what went through your mind when you received the invitation letter for our study?   - Why did you decide to take part in the study? - What expectations did you have of the study?   What do you think about research?   - Have you already participated in other studies? - Would you like to be involved in the development of research projects/research questions? |
| Previous hearing tests | And then you decided to do a hearing test for the study. Have you already had any kind of hearing test before? (e.g. military service, university, education, follow-up care)  If yes: Tell me about this hearing test.   - Who ordered the hearing test? Why? - When was it? (After completion of therapy?) - Where did the hearing test take place? - How did you make the appointment? - What was the hearing test like? - What was the result? - How do you remember the hearing test? - How much effort was the test for you? |
| Hearing problems | If hearing problems are known: What is it like for you to live with hearing problems?   - What is it like to know that you have hearing problems? - Does the issue come up often? - When do you notice it most? - What measures have been taken? |
| Hearing test at the hearing aid shop | Now I would like to know about your experience with the hearing test at the hearing aid shop.  Tell me about the hearing test at hearing aid shop.   - How come you made the appointment there? - How long did you have to wait for an appointment? - How did you travel to the branch? How long? - How much effort was involved for you? - What was the hearing test like? - What information did you receive from the acoustician? - How did you feel during the test? - Did you think about the possible test result? - How was it for you to talk to the acoustician about your hearing compared to a doctor? |
| After the hearing test | How did you feel after the hearing test?  How did you react when you received the result of your hearing test?   - How were you informed of the result? - How did you feel about the result afterwards?   Did the hearing test change anything in your everyday life?   - Did/will you discuss the result with someone?   - If yes: With whom?   - If yes: What did they do/say?   - If no: Do you still plan to do this? |
| Advantages & disadvantages of hearing test options | If you now compare this and your previous hearing test experience (if applicable): What are the differences between the two tests?   - What are the advantages/disadvantages of both?   What are the differences between an acoustician in the hearing aid shop and a doctor who carries out the hearing test?   - What are the advantages/disadvantages on both sides?   If you had to have a hearing test regularly, what would be most important to you during the visit?  Where would you go for a hearing test in the future?   - Why there? - What are the advantages for you personally of doing it at this location?   What are the disadvantages for you personally if you do it somewhere else? |
| Follow-up care setting | I would now like to talk to you about follow-up care after a cancer diagnosis. By this I mean examinations, such as blood tests, to detect any effects of your treatment or illness at an early stage after treatment has been completed.  Do you go for a follow-up examination or another regular check-up? (e.g. annual visit to the general practitioner)  What do they examine? |
| Transition to adult follow-up care | What was the transition from pediatric to adult follow-up care like for you?  Can you remember the information you received about follow-up care?   - What was particularly emphasized? - Who did you receive the information from? - Was the information verbal/written? - Would you have liked more information at the time? - Was hearing follow-up a topic? |
| Current follow-up care | What do you think about follow-up care today?   - What is the current importance of follow-up care in your life?   - Do you still go to follow-up examinations?     - Where?     - Why / why not?     - How does it work?     - Do they check your hearing?     - How does it make you feel?   What could be improved in follow-up care?   - What would you wish for new survivors? - What is already going well in terms of follow-up care? |
| End | Has your view of follow-up care changed as a result of the study?  What expectations did the study (not) fulfill?  What do you take away from this study?  Now I've almost finished my questions. Perhaps you would like to tell me something else that I haven't asked?  Do you have any questions for me or about the study? |
